# Changing Trends in Mechanical Circulatory Support Utilization and Outcomes in Patients Undergoing Percutaneous Coronary Interventions for Acute Coronary Syndrome Complicated with Cardiogenic Shock: Insights from a Nationwide Registry in Japan

**DOI:** 10.1101/2023.05.03.23289484

**Authors:** Yuji Nishimoto, Taku Inohara, Shun Kohsaka, Kenichi Sakakura, Tsutomu Kawai, Atsushi Kikuchi, Tetsuya Watanabe, Takahisa Yamada, Masatake Fukunami, Kyohei Yamaji, Hideki Ishii, Tetsuya Amano, Ken Kozuma, J-PCI Registry Investigators

**Affiliations:** Division of Cardiology, Osaka General Medical Center, Osaka, Japan; Department of Cardiology, Keio University School of Medicine, Tokyo, Japan; Division of Cardiovascular Medicine, Saitama Medical Center, Jichi Medical University, Saitama, Japan; Department of Cardiology, Kyoto University, Kyoto, Japan; Department of Cardiovascular Medicine, Gunma University Graduate School of Medicine, Gunma, Japan; Department of Cardiology, Aichi Medical University, Nagakute, Japan; Department of Cardiology, Teikyo University Hospital, Tokyo, Japan

**Author notes:** **Address for correspondence:** Yuji Nishimoto, MD, Division of Cardiology, Osaka General Medical Center, 3-1-56, Mandai-Higashi, Sumiyoshi-ku, Osaka 558-8558, Japan.

**Keywords:** cardiogenic shock, mechanical circulatory support, extracorporeal membrane oxygenation, intra-aortic balloon pumping, Impella, epidemiology

## Abstract

**Background:** Temporal trends in the management of acute coronary syndrome (ACS) complicated with cardiogenic shock (CS) after the revision of the guideline recommendations for intra-aortic balloon pump (IABP) use and the approval of the Impella require further investigation as their impact remains uncertain.

**Methods:** Using the Japanese Percutaneous Coronary Intervention (J-PCI) registry database from 2019 to 2021 (734,379 patients from 1,190 hospitals), we extracted 24,516 patients undergoing PCI for ACS complicated with CS. Of those, 12,171 patients (49.6%) used mechanical circulatory support (MCS) during the procedure. The patients were stratified into three groups: (i) IABP alone, (ii) Impella, and (iii) venoarterial extracorporeal membrane oxygenation (VA-ECMO); the VA-ECMO group was further stratified into (iiia) VA-ECMO alone, (iiib) VA-ECMO in combination with the IABP, and (iiic) VA-ECMO in combination with the Impella (ECPella). The quarterly prevalence and outcomes were reported.

**Results:** During the study period, there were notable changes in the prevalence of different MCS modalities and their associated outcomes. The use of an IABP alone and VA-ECMO decreased significantly from 63.5% and 34.4% in the first quarter of 2019 to 58.3% and 33.0% in the fourth quarter of 2021, respectively (P for trend = 0.01 and 0.02, respectively). Among the subset of patients who required VA-ECMO (*n* = 4,245), the use of VA-ECMO in combination with the IABP decreased significantly from 78.7% to 67.3%, whereas the use of ECPella increased significantly from 4.2% to 17.0% (P for trend <0.001 for both). There was no significant change in the use of VA-ECMO alone. In-hospital mortality decreased significantly over time in both the overall population of patients requiring MCS and those requiring VA-ECMO (P for trend = 0.004 and <0.001, respectively).

**Conclusions:** In conclusion, our study revealed significant changes in the use of different MCS modalities and associated outcomes in ACS complicated with CS, highlighting the evolving patterns of MCS utilization during the study period.

## Introduction

Cardiogenic shock (CS) is the leading cause of death in acute myocardial infarction with early mortality at 30–50% over the past two decades,^1–6^ despite the widespread implementation of early revascularization and advances in critical care. In addition, the incidence of CS has increased especially in the older adults and patients with non-ST-elevation myocardial infarction (NSTEMI).

In recent years, guidelines in the United States (2013), Europe (2014), and Japan (2018) have downgraded the recommendations for the use of an intra-aortic balloon pump (IABP),^7–9^ due to the findings from previous studies that showed no survival benefit of IABP use for patients with acute myocardial infarction irrespective of concomitant CS.^10–12^ Consequently, the use of an IABP has declined in Europe and the United States, while the use of an Impella, a new percutaneous ventricular assist device, is on the rise.^6,13^ Since the availability of the Impella in Japan starting from October 2017, where IABPs were predominantly used,^14^ it remains uncertain how these significant changes have influenced the utilization of mechanical circulatory support (MCS) in patients with CS and their clinical outcomes, especially in the context of Japan being a super-aged society.

It is essential to capture the trends in the management of acute coronary syndrome (ACS) complicated with CS in light of the changes in the guideline recommendations, and approval of the new MCS device is crucial to ascertain the current status of CS practice and to establish evidence-based medicine in future practice. Accordingly, the present study aimed to describe the prevalence of MCS utilization and outcomes in patients undergoing percutaneous coronary intervention (PCI) for ACS complicated with CS and their trends over time, using a nationwide PCI registry database in Japan.

## Methods

### Data source

We used the Japanese Percutaneous Coronary Intervention (J-PCI) registry database, which contained clinical data on patients undergoing PCI from 1,190 hospitals, including all hospitals using the Impella, and covered approximately 90% of all PCI procedures in Japan.^15–17^ The J-PCI registry is administrated by the Japanese Association of Cardiovascular Intervention and Therapeutics (CVIT) and its registration is mandatory for the application of board certification and renewal, ensuring a high level of data completeness. The accuracy of the submitted data is maintained by data auditing (20 hospitals annually), which is operated by members of the CVIT subcommittee. The database includes the patient background, clinical presentation, angiographic and procedural information, and in-hospital outcomes.

### Design and Ethical statement

This was a retrospective cohort study using a nationwide registry database, and the study conformed to the RECORD reporting guidelines.^18^ This study was conducted in accordance with the amended Declaration of Helsinki and was approved by the Institutional Review Board Committee of the Network for Promotion of Clinical Studies, a nonprofit organization affiliated with Osaka University Graduate School of Medicine, Osaka, Japan. Because the data were anonymous, the Institutional Review Board waived the requirement for informed consent.

### Study population

Using the data from January 2019 to December 2021, we included patients undergoing PCI from 1,190 hospitals (*n* = 734,379) (**Figure 1**). We excluded the patients with missing information of the outcomes (*n* = 50) and those outside the age range of 20 to 100 years (*n* = 106). We also excluded patients who underwent PCI with a clinical presentation other than ACS (*n* = 45,904), in an elective setting (*n* = 78,280), and without CS (*n* = 178,523). Of 24,516 patients undergoing PCI for ACS complicated with CS from 1,072 hospitals, we also excluded patients who underwent PCI without MCS (*n* = 12,101) and those with MCS other than IABP / venoarterial extracorporeal membrane oxygenation (VA-ECMO) / Impella (*n* = 244). We ultimately identified 12,171 patients undergoing PCI for ACS complicated with CS under MCS from 937 hospitals.

**Figure 1.**
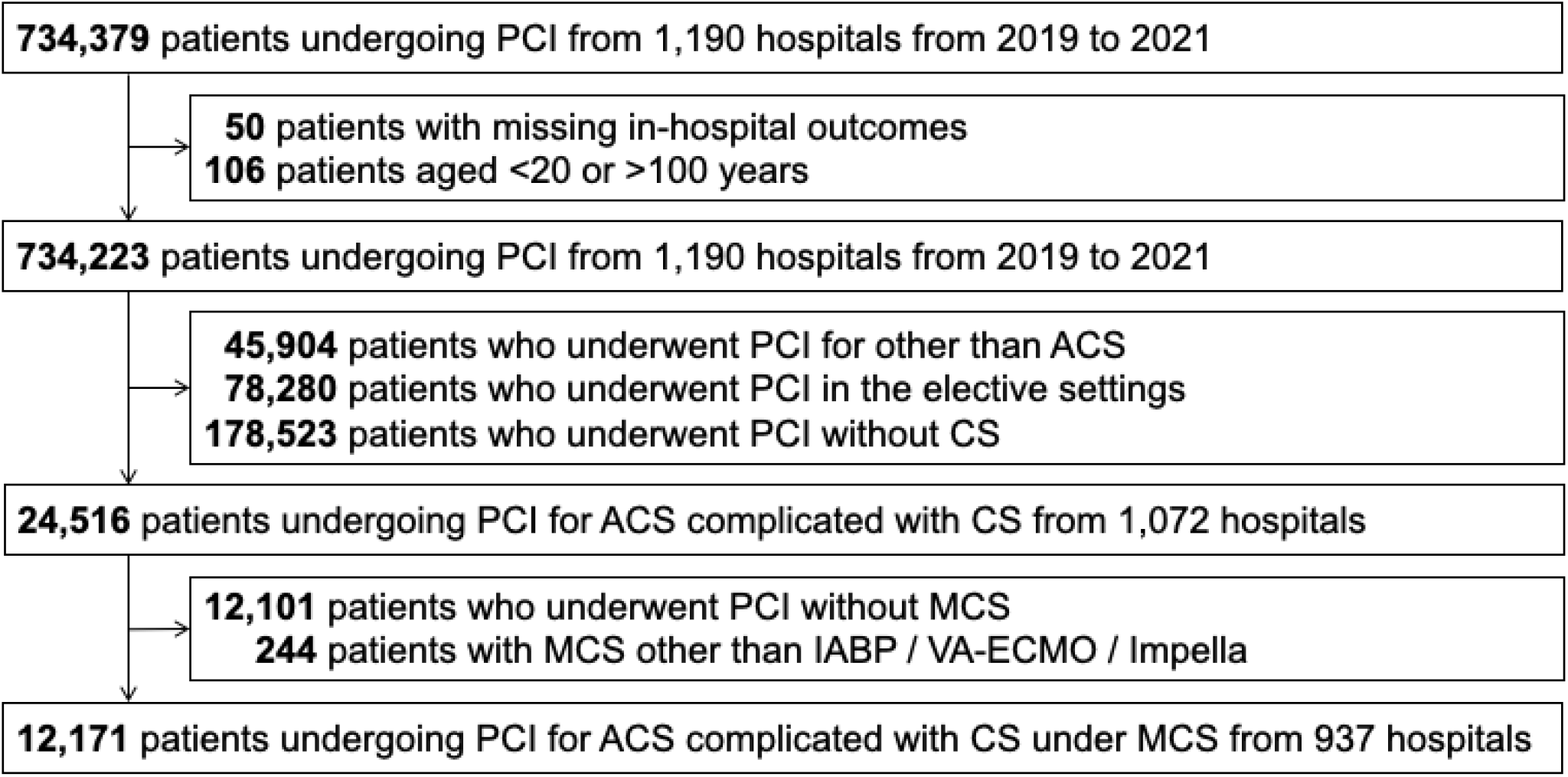
Patient flowchart ACS, acute coronary syndrome; CS, cardiogenic shock; IABP, intra-aortic balloon pump; PCI, percutaneous coronary intervention; VA-ECMO, venoarterial extracorporeal membrane oxygenation

The eligible patients were stratified into three groups according to the MCS modalities: (i) IABP alone, (ii) Impella, and (iii) VA-ECMO. The VA-ECMO group was further stratified into three groups according to the concomitant MCS devices: (iiia) VA-ECMO alone, (iiib) VA-ECMO + IABP, and (iiic) VA-ECMO + Impella (ECPella). Given that the presence of patients receiving these modalities overlapped (**Supplementary Table S1**), we categorized the groups according to the following hierarchy: VA-ECMO outweighs the Impella, and the Impella outweighs IABP.^19^

### Variables and outcomes

The variables included the age, sex, medical history (prior myocardial infarction, heart failure, PCI, or coronary artery bypass grafting), comorbidities (diabetes mellitus, hypertension, dyslipidemia, current smoker, chronic kidney disease, hemodialysis, chronic obstructive pulmonary disease, or peripheral artery disease), clinical presentation (ST-elevation myocardial infarction [STEMI], NSTEMI, heart failure within 24 hours, or cardiac arrest within 24 hours), laboratory data (hemoglobin or creatinine), number of diseased vessels, target lesions, access site, door-to-balloon time, timing of MCS utilization, pretreatment medication status regarding antiplatelet agents or oral anticoagulants, and treatment devices (non-stent, drug-eluting stents, bare metal stents, aspiration, or distal protection).

The definitions of the variables were described elsewhere.^20^ In brief, heart failure was defined as symptoms of heart failure within 24 hours before the PCI procedure, including dyspnea on mild activity, orthopnea, body fluid retention, moist rales, neck vein distention, and pulmonary oedema, which were equivalent to congestive heart failure of a New York Heart Association functional classification Class IV. CS was defined as a sustained episode of systolic blood pressure of <80 mmHg, cardiac index of <1.8 L/min/m^2^ determined to be secondary to cardiac dysfunction, and/or the requirement for a parenteral inotropic or vasopressor agent or MCS to maintain the blood pressure and cardiac index above the specified levels. Chronic kidney disease in this registry was defined as the presence of proteinuria, a serum creatinine of ≥1.3 mg/dL, or an estimated glomerular filtration rate of ≤60 mL/min/1.73 m^2^, according to the guidelines from the Japanese Society of Nephrology.^21^ Preoperative and intraoperative MCS utilization was defined when the MCS was introduced before entry and exit from the catheterization laboratory, respectively.^19^

The primary outcome was in-hospital mortality. The secondary outcomes were procedural complications (cardiac tamponade, heart failure or shock status, definite stent thrombosis according to the Academic Research Consortium definition, emergent surgery, or bleeding requiring blood transfusion), procedural time, amount of contrast media, or TIMI (Thrombolysis in Myocardial Infarction) flow grade 3 at the end of the procedure.

#### Statistical analysis

Continuous variables were presented as the mean and standard deviation, and categorical variables were presented as the number and percentage. Continuous variables were compared using a one-way analysis of variance, and categorical variables were compared using the chi-squared test. To evaluate the trends by quarter within each year, we performed Cochran-Armitage tests for the binary variables. The trends in the primary outcome were assessed among the overall patients requiring MCS and those requiring VA-ECMO. The trends in the prevalence of each MCS device were also assessed. We considered all reported p-values as two-sided and a p<0.05 as statistically significant. All analyses were performed using R software version 4.0.5 (R Foundation for Statistical Computing, Vienna, Austria).

## Results

### Baseline characteristics and procedural details

Of the 12,171 eligible patients, 7,304 (60.0%) were identified as the IABP alone group, 622 (5.1%) as the Impella group, and the remaining 4,245 (34.9%) as the VA-ECMO group (**Table 1**). Of the VA-ECMO group, 728 patients (17.1%) were identified as the VA-ECMO alone group, 3,027 (71.3%) as the VA-ECMO + IABP group, and remaining 480 (11.3%) as the ECPella group. The mean age of the overall patient cohort was 71 years, with 23.0% being women, and 77.0% presenting with STEMI. Among the different MCS modalities, the patients in the IABP alone group were the oldest, while those in the ECPella group were the youngest. The prevalence of patients with STEMI was highest in the ECPella group and lowest in the VA-ECMO alone group, while those with NSTEMI were highest in the VA-ECMO alone group and lowest in the ECPella group. The prevalence of cardiac arrest within the 24 hours prior to the PCI procedure varied among the different MCS modalities, with the highest rate observed in the VA-ECMO alone group (86.5%) and the lowest rate in the Impella group (28.6%), with the IABP group at 32.6%, the VA-ECMO + IABP group at 81.8%, and the ECPella group at 76.2%. The Impella group had the highest frequency of 3-vessel disease and the lowest frequency of culprit lesions in the right coronary artery. Antiplatelet therapy and drug-eluting stents were most frequently used in the Impella group, while they were least frequently used in the VA-ECMO alone group, which instead had a higher utilization of non-stent strategies.

**Table 1.** Baseline characteristics and procedural details.

|  | Overall | IABP alone | Impella | VA-ECMO |  |  |
| --- | --- | --- | --- | --- | --- | --- |
|  |  |  |  | VA-ECMO alone | VA-ECMO + IABP | ECPella |
|  | (*n* = 12,171) | (*n* = 7,304) | (*n* = 622) | (*n* = 728) | (*n* = 3,037) | (*n* = 480) |
| Age, years, mean (SD) | 70.79 (12.46) | 73.11 (12.06) | 70.13 (12.01) | 68.39 (12.60) | 66.62 (12.12) | 66.40 (12.14) |
| Women, *n* (%) | 2,803 (23.0) | 1,946 (26.6) | 120 (19.3) | 153 (21.0) | 503 (16.6) | 81 (16.9) |
| Medical history, *n* (%) |  |  |  |  |  |  |
| Prior myocardial infarction | 1,872 (15.4) | 1,172 (16.0) | 93 (15.0) | 96 (13.2) | 448 (14.8) | 63 (13.1) |
| Prior heart failure | 2,092 (17.2) | 1,339 (18.3) | 113 (18.2) | 120 (16.5) | 445 (14.7) | 75 (15.6) |
| Prior percutaneous coronary intervention | 2,245 (18.4) | 1,384 (18.9) | 107 (17.2) | 148 (20.3) | 538 (17.7) | 68 (14.2) |
| Prior coronary artery bypass grafting | 333 (2.7) | 193 (2.6) | 12 (1.9) | 36 (4.9) | 82 (2.7) | 10 (2.1) |
| Comorbidities, *n* (%) |  |  |  |  |  |  |
| Diabetes mellitus | 5,372 (44.1) | 3,281 (44.9) | 277 (44.5) | 289 (39.7) | 1,327 (43.7) | 198 (41.2) |
| Hypertension | 7,882 (64.8) | 4,927 (67.5) | 412 (66.2) | 443 (60.9) | 1,829 (60.2) | 271 (56.5) |
| Dyslipidemia | 6,113 (50.2) | 3,871 (53.0) | 358 (57.6) | 302 (41.5) | 1,364 (44.9) | 218 (45.4) |
| Current smoker | 3,954 (32.5) | 2,302 (31.5) | 213 (34.2) | 221 (30.4) | 1,046 (34.4) | 172 (35.8) |
| Chronic kidney disease | 4,182 (34.4) | 2,636 (36.1) | 235 (37.8) | 234 (32.1) | 912 (30.0) | 165 (34.4) |
| Hemodialysis | 643 (5.3) | 408 (5.6) | 21 (3.4) | 49 (6.7) | 146 (4.8) | 19 (4.0) |
| Chronic obstructive pulmonary disease | 404 (3.3) | 244 (3.3) | 26 (4.2) | 24 (3.3) | 95 (3.1) | 15 (3.1) |
| Peripheral artery disease | 819 (6.7) | 499 (6.8) | 44 (7.1) | 83 (11.4) | 171 (5.6) | 22 (4.6) |
| Clinical presentation, *n* (%) |  |  |  |  |  |  |
| ST-elevation myocardial infarction | 9,371 (77.0) | 5,618 (76.9) | 475 (76.4) | 516 (70.9) | 2,368 (78.0) | 394 (82.1) |
| Non-ST-elevation myocardial infarction | 2,109 (17.3) | 1,246 (17.1) | 106 (17.0) | 173 (23.8) | 519 (17.1) | 65 (13.5) |
| Heart failure within 24 hours | 7,781 (63.9) | 4,577 (62.7) | 427 (68.6) | 431 (59.2) | 2,007 (66.1) | 339 (70.6) |
| Cardiac arrest within 24 hours | 6,037 (49.6) | 2,379 (32.6) | 178 (28.6) | 630 (86.5) | 2,484 (81.8) | 366 (76.2) |
| Laboratory data, mean (SD) |  |  |  |  |  |  |
| Hemoglobin, g/dL | 12.95 (2.53) | 12.90 (2.47) | 13.18 (2.41) | 12.52 (2.66) | 13.09 (2.64) | 13.10 (2.51) |
| Creatinine, mg/dL | 1.68 (1.83) | 1.70 (1.84) | 1.45 (1.26) | 1.79 (2.00) | 1.69 (1.88) | 1.60 (1.64) |
| Number of diseased vessels, *n* (%) |  |  |  |  |  |  |
| 1-vessel disease | 4,862 (39.9) | 2,829 (38.7) | 215 (34.6) | 336 (46.2) | 1,289 (42.4) | 193 (40.2) |
| 2-vessel disease | 3,429 (28.2) | 2,163 (29.6) | 164 (26.4) | 183 (25.1) | 788 (25.9) | 131 (27.3) |
| 3-vessel disease | 3,284 (27.0) | 2,049 (28.1) | 207 (33.3) | 148 (20.3) | 764 (25.2) | 116 (24.2) |
| Left main disease | 2,212 (18.2) | 1,135 (15.5) | 156 (25.1) | 143 (19.6) | 642 (21.1) | 136 (28.3) |
| Target lesions, *n* (%) |  |  |  |  |  |  |
| Right coronary artery | 4,429 (36.4) | 2,797 (38.3) | 175 (28.1) | 258 (35.4) | 1,059 (34.9) | 140 (29.2) |
| Left main trunk or left anterior descending artery | 8,380 (68.9) | 4,800 (65.7) | 510 (82.0) | 485 (66.6) | 2,191 (72.1) | 394 (82.1) |
| Left circumflex artery | 2,951 (24.2) | 1,747 (23.9) | 177 (28.5) | 144 (19.8) | 744 (24.5) | 139 (29.0) |
| Access site, *n* (%) |  |  |  |  |  |  |
| Radial | 3,917 (32.2) | 2,677 (36.7) | 290 (46.6) | 161 (22.1) | 662 (21.8) | 127 (26.5) |
| Femoral | 7,648 (62.8) | 4,355 (59.6) | 306 (49.2) | 512 (70.3) | 2,156 (71.0) | 319 (66.5) |
| Other | 606 (5.0) | 272 (3.7) | 26 (4.2) | 55 (7.6) | 219 (7.2) | 34 (7.1) |
| Door-to-balloon time, min, mean (SD) (*n* = 7,762) | 95.60 (56.51) | 94.34 (56.30) | 101.05 (54.42) | 106.16 (62.08) | 95.80 (57.04) | 93.17 (50.00) |
| Medication, *n* (%) |  |  |  |  |  |  |
| Preprocedural antiplatelet therapy | 9,410 (77.3) | 5,871 (80.4) | 513 (82.5) | 472 (64.8) | 2,176 (71.6) | 378 (78.8) |
| Dual antiplatelet therapy | 8,104 (66.6) | 5,084 (69.6) | 436 (70.1) | 390 (53.6) | 1,884 (62.0) | 310 (64.6) |
| P2Y<sub>12</sub> receptor inhibitor | 8,409 (69.1) | 5,273 (72.2) | 459 (73.8) | 403 (55.4) | 1,952 (64.3) | 322 (67.1) |
| Preprocedural oral anticoagulants | 576 (4.7) | 373 (5.1) | 28 (4.5) | 35 (4.8) | 123 (4.1) | 17 (3.5) |
| Direct oral anticoagulants | 350 (2.9) | 232 (3.2) | 17 (2.7) | 18 (2.5) | 73 (2.4) | 10 (2.1) |
| Treatment, *n* (%) |  |  |  |  |  |  |
| Non-stent | 2,189 (18.0) | 1,368 (18.7) | 71 (11.4) | 182 (25.0) | 507 (16.7) | 61 (12.7) |
| Drug-eluting stents | 9,951 (81.8) | 5,912 (80.9) | 551 (88.6) | 544 (74.7) | 2,525 (83.1) | 419 (87.3) |
| Bare metal stents | 46 (0.4) | 33 (0.5) | 1 (0.2) | 2 (0.3) | 10 (0.3) | 0 (0.0) |
| Aspiration | 5,010 (41.2) | 3,134 (42.9) | 217 (34.9) | 219 (30.1) | 1,245 (41.0) | 195 (40.6) |
| Distal protection | 555 (4.6) | 362 (5.0) | 23 (3.7) | 17 (2.3) | 128 (4.2) | 25 (5.2) |

ECPella, combination of VA-ECMO and Impella; IABP, intra-aortic balloon pumping; SD, standard deviation; VA-ECMO, venoarterial extracorporeal membrane oxygenation

### In-hospital outcomes

**Table 2** shows that in-hospital mortality rates were highest in the VA-ECMO alone group (58.5%) and lowest in the Impella group (24.6%), with the IABP alone group at 26.1%, the VA-ECMO + IABP group at 55.7%, and the ECPella group at 46.9%. Bleeding requiring blood transfusions was most prevalent in the ECPella group (8.1% and 6.0% for access and non-access sites, respectively) and least prevalent in the IABP alone group (0.7% and 1.2%). The achievement of TIMI flow grade 3 at the end of the procedure was most prevalent in the ECPella group (98.1%) and least prevalent in the VA-ECMO alone group (93.0%), with the Impella group at 97.6%, the VA-ECMO + IABP group at 96.5%, and the IABP alone group at 95.7%.

**Table 2.** In-hospital outcomes.

|  | Overall | IABP alone | Impella | VA-ECMO |  |  |
| --- | --- | --- | --- | --- | --- | --- |
|  |  |  |  | VA-ECMO alone | VA-ECMO + IABP | ECPella |
|  | (n = 12,171) | (n = 7,304) | (n = 622) | (n = 728) | (n = 3,037) | (n = 480) |
| In-hospital mortality, n (%) | 4,405 (36.2) | 1,908 (26.1) | 153 (24.6) | 426 (58.5) | 1,693 (55.7) | 225 (46.9) |
| Procedural complications, n (%) | 2,112 (17.4) | 1,051 (14.4) | 117 (18.8) | 130 (17.9) | 690 (22.7) | 124 (25.8) |
| Cardiac tamponade | 100 (0.8) | 32 (0.4) | 4 (0.6) | 13 (1.8) | 41 (1.4) | 10 (2.1) |
| Heart failure or shock status | 1,641 (13.5) | 867 (11.9) | 71 (11.4) | 85 (11.7) | 544 (17.9) | 74 (15.4) |
| Stent thrombosis | 110 (0.9) | 74 (1.0) | 1 (0.2) | 9 (1.2) | 24 (0.8) | 2 (0.4) |
| Emergent surgery | 115 (0.9) | 55 (0.8) | 6 (1.0) | 9 (1.2) | 41 (1.4) | 4 (0.8) |
| Bleeding requiring blood transfusions | 560 (4.6) | 138 (1.9) | 55 (8.8) | 57 (7.8) | 248 (8.2) | 62 (12.9) |
| Access site | 261 (2.1) | 53 (0.7) | 26 (4.2) | 24 (3.3) | 119 (3.9) | 39 (8.1) |
| Non-access site | 319 (2.6) | 86 (1.2) | 32 (5.1) | 34 (4.7) | 138 (4.5) | 29 (6.0) |
| Procedural time, min, mean (SD) | 151.38 (74.03) | 139.87 (65.03) | 178.82 (81.21) | 146.05 (80.93) | 167.37 (82.39) | 195.20 (81.03) |
| Amount of contrast media, ml, mean (SD) | 148.54 (73.69) | 148.15 (72.31) | 162.56 (79.67) | 140.73 (80.11) | 148.11 (74.36) | 149.38 (69.80) |
| TIMI flow grade 3 at the end of procedure, n (%) | 11,673 (95.9) | 6,987 (95.7) | 607 (97.6) | 677 (93.0) | 2,931 (96.5) | 471 (98.1) |

ECPella, combination of VA-ECMO and Impella; IABP, intra-aortic balloon pumping; SD, standard deviation; TIMI, Thrombolysis in Myocardial Infarction; VA-ECMO, venoarterial extracorporeal membrane oxygenation

### Trends in the MCS utilization and outcomes

Figure 2. and **Supplementary Table S2** show that the use of IABP alone and VA-ECMO significantly decreased from 63.5% and 34.4% in 2019 Q1 to 58.3% and 33.0% in 2021 Q4, respectively (P for trend = 0.01 and 0.02, respectively). In contrast, the use of an Impella significantly increased from 2.1% in 2019 Q1 to 8.7% in 2021 Q4 (P for trend <0.001). Among the overall 12,171 patients requiring MCS, in-hospital mortality significantly decreased from 36.6% in 2019 Q1 to 32.1% in 2021 Q4 (P for trend = 0.004).

**Figure 2.**
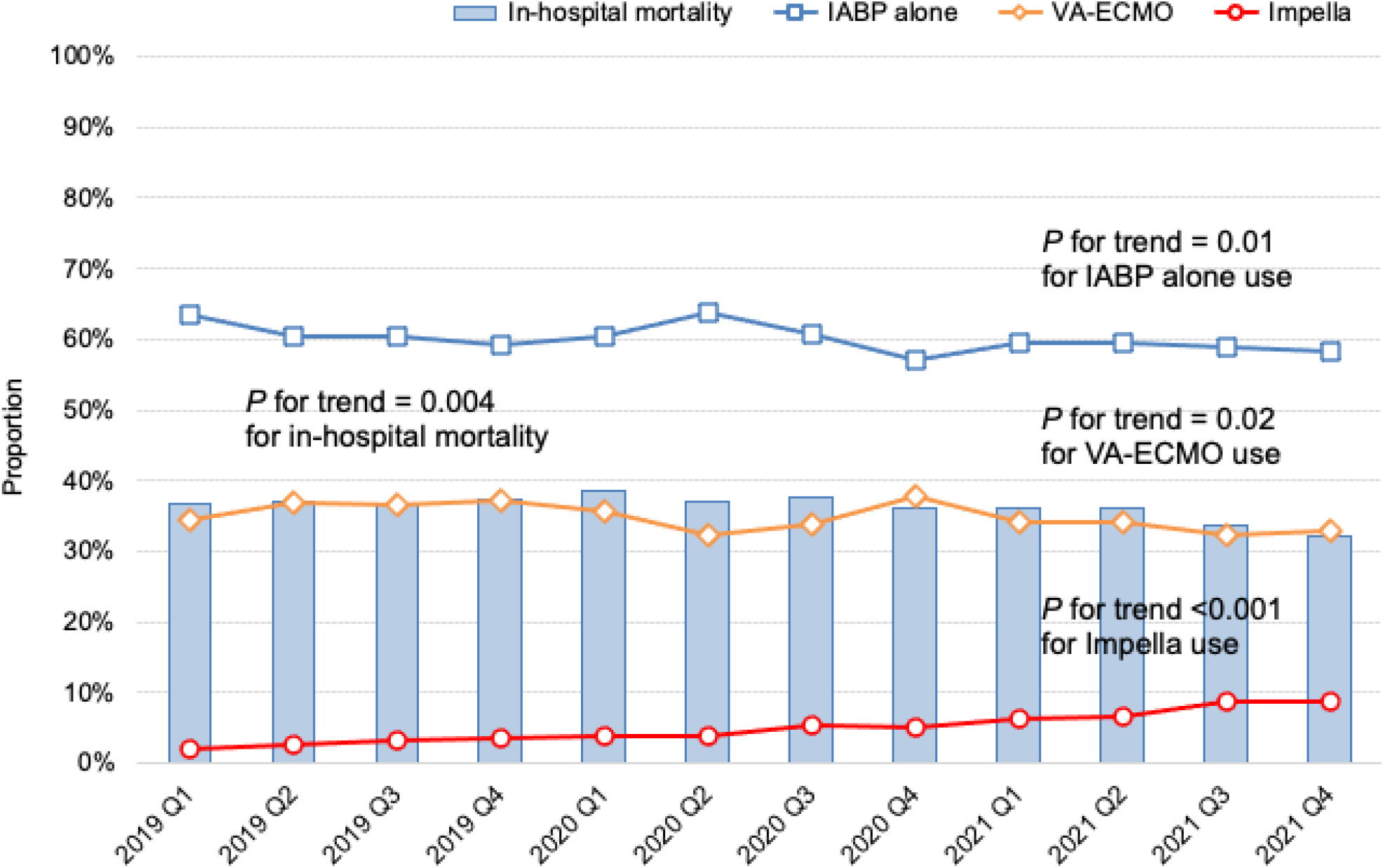
Quarterly proportion of in-hospital mortality and prevalence of IABP alone, Impella, and VA-ECMO use in the overall patients undergoing percutaneous coronary intervention for acute coronary syndrome complicated with cardiogenic shock under mechanical circulatory support IABP, intra-aortic balloon pump; VA-ECMO, venoarterial extracorporeal membrane oxygenation

Figure 3. and **Supplementary Table S3** reveal that among 4,245 patients requiring VA-ECMO, the use of VA-ECMO + IABP significantly decreased from 78.7% in 2019 Q1 to 67.3% in 2021 Q4 (P for trend <0.001). In contrast, the use of ECPella significantly increased from 4.2% in 2019 Q1 to 17.0% in 2021 Q4 (P for trend <0.001). There was no significant change in the prevalence of VA-ECMO alone (P for trend = 0.15). In-hospital mortality significantly decreased from 59.8% in 2019 Q1 to 50.0% in 2021 Q4 (P for trend <0.001).

**Figure 3.**
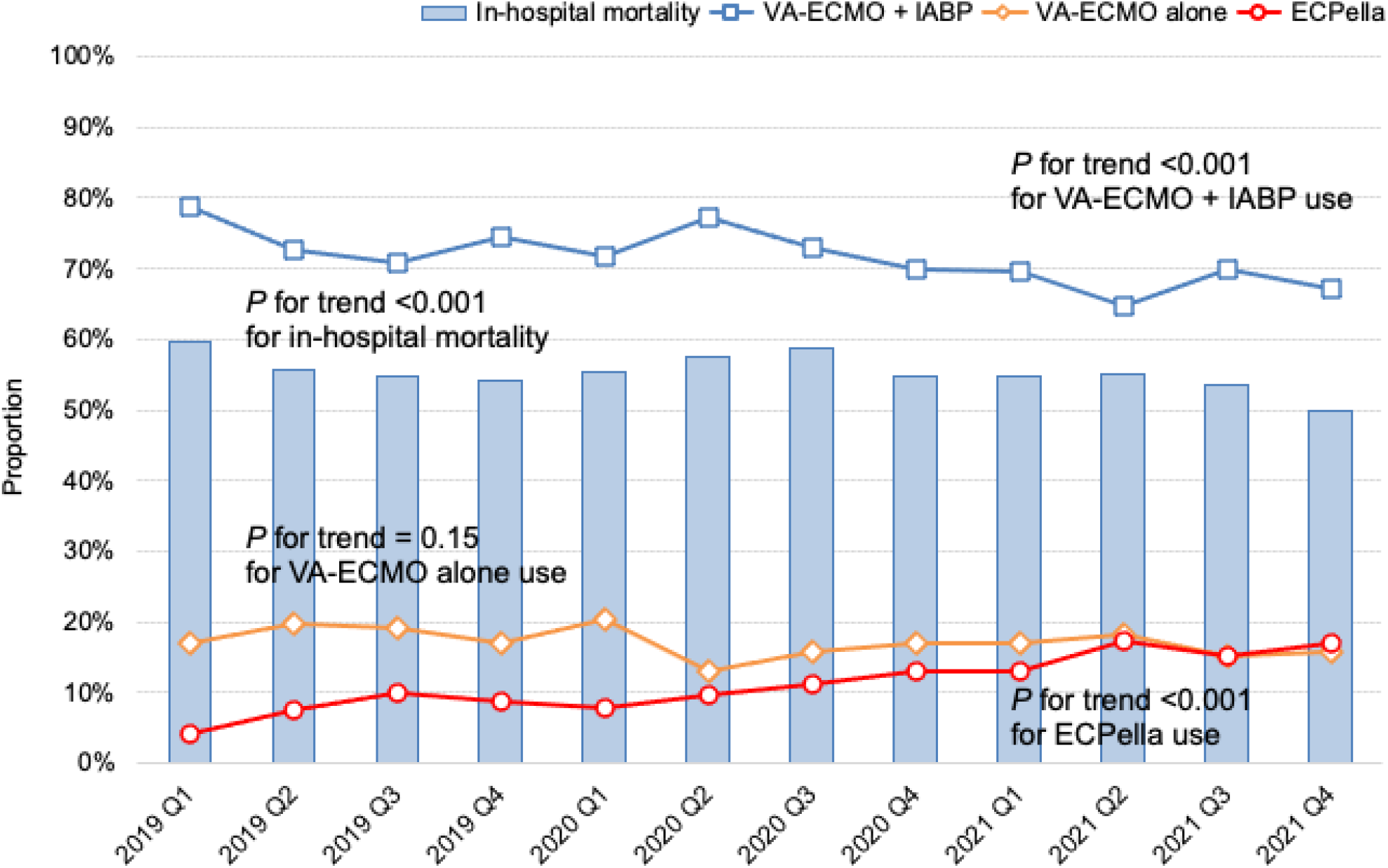
Quarterly proportion of in-hospital mortality and prevalence of VA-ECMO alone, VA-ECMO + IABP, and ECPella use in the patients undergoing percutaneous coronary intervention for acute coronary syndrome complicated with cardiogenic shock under VA-ECMO ECPella, combination of VA-ECMO and Impella; IABP, intra-aortic balloon pump; VA-ECMO, venoarterial extracorporeal membrane oxygenation

## Discussion

To the best of our knowledge, this is the first nationwide study on time-trends of MCS utilization and the outcomes in patients undergoing PCI for ACS complicated with CS in Japan. Among the patients requiring MCS, there were decreasing trends in IABP alone use and VA-ECMO use, while there was an increasing trend in Impella use from 2019 to 2021. Among the patients requiring VA-ECMO, there was an increasing trend in ECPella use from 2019 to 2021. There was a decreasing trend in in-hospital mortality in both the overall patients requiring MCS and those requiring VA-ECMO, potentially highlighting the positive impact of the changes in the patterns of MCS utilization during the study period.

Compared to the previous studies from Europe and the United States,^13,23–27^ the present study showed similar temporal trends of a decreasing IABP use and increasing Impella use, but the degree of those changes substantially differed. In Europe, the use of an IABP drastically decreased and was mostly replaced by an Impella,^25,27^ while in the United States, IABPs have been predominantly used despite a greatly increasing trend of Impella use.^24,26^ The present study from the Japanese nationwide registry revealed that the use of an IABP had been predominant over time and the use of an Impella remained minor. This result may be attributed to the very high penetration of the IABP among hospitals performing PCI in Japan, the lower profile of the IABP catheters as compared to the Impella and ECMO catheters, and its frequent use in elective high-risk PCI.^19^ In addition, the Impella was only available for less than 200 hospitals in 2021 and has been limited to use only for patients with CS, not for those undergoing high-risk PCI.^28,29^

In the present study, in-hospital mortality in the overall patients requiring MCS was 36.2%, which was consistent with previous studies.^30,31^ However, data on temporal trends in in-hospital mortality remains scarce. The present study revealed a decreasing trend in in-hospital mortality in the overall patients requiring MCS, which may be attributed to the decreasing prevalence of patients requiring VA-ECMO who had an approximately twice higher in-hospital mortality as compared to those requiring other than VA-ECMO in the present study. In-hospital mortality in the patients requiring VA-ECMO was 46.9–58.5% with a decreasing trend, which was almost consistent with those in the previous study.^32^ The increase in the use of ECPella was about twice as large as that of the Impella (12.8% versus 6.6%), which might be partly due to the expectation of left ventricular unloading. Further investigation is required as to whether left ventricular unloading by the concomitant use of an Impella with VA-ECMO improves the outcome as compared to that by the concomitant use of an IABP with VA-ECMO.

Our study findings have several important clinical implications. First, the downgrade of the guideline recommendations for IABP use and approval of the Impella have likely influenced the pattern of MCS utilization, particularly with an increasing use of ECPella, albeit not drastically, in Japan where the Impella has only been available in limited hospitals. Second, while there was a decreasing trend in in-hospital mortality in both the overall patients requiring MCS and those requiring VA-ECMO, it remained high, especially in patients requiring VA-ECMO. Achieving further mortality reductions, left ventricular unloading by the concomitant use of an IABP or Impella with VA-ECMO might be a key, but further investigation is required.

The present study had several strengths. Firstly, it utilized one of the largest databases from more than 1,000 hospitals, including all hospitals using an Impella, covering approximately 90% of all PCI procedures in Japan.^34^ Secondly, the database included complete data on in-hospital mortality, enhancing the robustness of the findings. However, there were also several limitations to our study. Firstly, our definition of CS was based on historical studies such as the SHOCK trial,^35^ and detailed clinical information, such as the vital signs and laboratory tests, were not available in the database. Additionally, the groups classified based on the MCS modalities were determined using information available at the time of the PCI procedure. Any MCS modalities that were added after the PCI procedure were not considered in the present analysis. Potential misclassifications and bias could arise due to these factors. Moreover, since the decision on MCS utilization was left to the discretion of individual clinicians, the findings may be subject to variability. Lastly, the present study was observational in nature, and causality between the use of each MCS device and in-hospital mortality cannot be inferred.

## Conclusions

The present study using a nationwide PCI registry database showed significant temporal changes in the patterns of MCS utilization and clinical outcomes in patients undergoing PCI for ACS complicated with CS. Among the overall patients requiring MCS, the use of IABP alone and VA-ECMO decreased, while the use of an Impella increased. Among the patients requiring VA-ECMO, the use of VA-ECMO + IABP decreased, while the use of ECPella increased. There was a decreasing trend in in-hospital mortality in the overall patients requiring MCS as well as those requiring VA-ECMO. These findings highlight the dynamic nature of MCS utilization and its impact on clinical outcomes in this high-risk population. Further research is warranted to explore the optimal MCS strategies to improve the patient outcomes in ACS complicated with CS.

## Data Availability

The data, analytic methods, and study materials will not be made publicly available to other researchers for the purpose of reproducing the results or replicating the procedure.

## Acknowledgments

We would like to express our gratitude to Mr. John Martin for his grammatical assistance.

## Sources of Funding

This work was supported by the Japanese Association of Cardiovascular Intervention.

## Disclosures

### Ethics approval and consent to participate

This study was performed in accordance with the amended Declaration of Helsinki, and was approved by the Institutional Review Board Committee of the Network for Promotion of Clinical Studies, a nonprofit organization affiliated with Osaka University Graduate School of Medicine, Osaka, Japan.

### Consent for publication

The review board waived the requirement for informed consent because of the anonymous nature of the data. No information describing the individual patients, hospitals, or treating physicians was obtained.

### Competing interests

Dr. Nishimoto received lecture fees from Bayer Healthcare, Bristol-Myers Squibb, Pfizer, and Daiichi-Sankyo. Dr. Kohsaka reported investigator-initiated grant from Novartis and Daiichi Sankyo, and personal fees from Bristol-Myers Squibb. Dr. Sakakura received lecture fees from Abbott Vascular, Boston Scientific, Medtronic Cardiovascular, Terumo, OrbusNeich, Japan Lifeline, Kaneka, and Nipro. Dr. Ishii received lecture fees from Astellas Pharma, AstraZeneca, Bayer, Bristol-Myers Squibb, Chugai Pharma, Daiichi Sankyo, Kowa, Novartis Pharma, Nippon Boehringer Ingelheim, Otsuka Pharma, Pfizer, Mochida Pharma, and Tanabe Mitsubishi. Dr. Amano received lecture fees from Astellas Pharma, AstraZeneca, Bayer, Daiichi Sankyo, and Bristol-Myers Squibb. The remaining authors have no conflicts of interest to declare.

### Authors’ contributions

**Yuji Nishimoto**: Conceptualization, Software, Investigation, Writing - Original Draft; **Taku Inohara**: Conceptualization, Data curation, Software, Formal analysis, Investigation; **Shun Kohsaka**: Conceptualization, Investigation, Writing - Review and Editing; **Kenichi Sakakura**: Writing - Review and Editing, Supervision; **Tsutomu Kawai**: Investigation, Supervision; **Atushi Kikuchi:** Investigation, Supervision; **Tetsuya Watanabe**: Investigation, Resources, Supervision; **Takahisa Yamada**: Investigation, Resources, Supervision; **Masatake Fukunami**: Writing - Review and Editing, Supervision; **Kyohei Yamaji**: Data curation, Software, Supervision; **Hideki Ishii**: Writing - Review and Editing, Supervision; **Tetsuya Amano**: Writing - Review and Editing, Supervision; **Ken Kozuma**: Writing - Review and Editing, Supervision;. All authors read and approved the final manuscript.

## Notes

### Clinical Trial

N/A

